# Blood pressure variability is an independent predictor of mortality in hypertensive patients aged 80 years and older, based on long-term ambulatory blood pressure monitoring

**DOI:** 10.64898/2026.03.26.26349458

**Authors:** Mengying Zeng, Mengjun Jiang, Yuchen Zhu, Yuanyuan Shang, Jingwen Shi, Yabing Wang, Ying Sun

## Abstract

1.

**Background:** Increasing evidence suggests that blood pressure variability (BPV) may offer prognostic value beyond average blood pressure levels. However, data on the association between BPV of ambulatory blood pressure monitoring and mortality in patients aged 80 and older are limited. This study aimed to investigate the relationship between BPV and all-cause mortality in this population.

**Methods:** A total of 5,838 ABPM records from the Geriatrics Department of Beijing Friendship Hospital, collected between October 12, 2018, and June 9, 2025, were analyzed. Patients were divided into death and non-death groups. Subgroup analyses were performed based on the number of completed ABPM sessions. Cox proportional hazards models assessed the associations between BPV and mortality. Kaplan–Meier analysis and log-rank tests were used to compare survival across groups.

**Results:** A median follow-up of 32.0 months included 727 hypertensive patients aged ≥80 years. Multivariable cox regression and kaplan–meier analyses showed that the reverse-dipper blood pressure pattern was significantly associated with increased mortality. While short-term BPV was not linked to mortality, greater long-term variability in nighttime SBP and daytime DBP was significantly associated with higher mortality.

**Conclusion:** Among individuals aged 80 and older, those with a reverse-dipper pattern and higher long-term BPV had a significantly higher mortality risk, despite achieving recommended blood pressure targets.

## 1. Introduction

With the arrival of an aging society, the number of elderly hypertension patients (≥80 years old) is gradually increasing. Based on the findings from the 2012-2015 National Hypertension Stratified Multistage Random Sampling Cross-sectional Survey, the prevalence of hypertension among elderly individuals in China is 60.27%(1). In recent years, studies have found that blood pressure variability (BPV), an indicator of the extent of blood pressure fluctuations, is more common in hypertensive patients, particularly notable in the elderly population(2).In elderly hypertensive patients, BPV manifests in various clinical forms, including circadian rhythm disorders (such as non-dipper or reverse dipper blood pressure patterns), postural blood pressure fluctuations (such as orthostatic hypotension or hypertension), and postprandial hypotension. These BPV patterns are closely associated with a range of adverse cardiovascular and cerebrovascular events and have a serious impact on the patient’s long-term prognosis. Studies have shown that 24-hour or short-term blood pressure variability is negatively correlated with cardiovascular risk, independent of the 24-hour average blood pressure value(3). The Kailuan study in China indicated that the highest quintile group of systolic blood pressure variability had a 37% and 18% increased risk of all-cause mortality and cardiovascular events, respectively, compared to the lowest quintile group(4). Although existing studies have demonstrated the clinical value of blood pressure variability in hypertensive patients, particularly its relationship with cardiovascular event risk, there is still limited systematic research on BPV in elderly hypertensive populations. Most research focuses on the impact of antihypertensive drug treatment regimens and different treatment approaches on blood pressure fluctuations or only addresses the control of average blood pressure, without in-depth exploration of the quantifiable indicators and manifestations of BPV on all-cause mortality.

In summary, the lack of BPV for elderly have prompted us to conduct this study. We aim to investigate the BPV derived from multiple ABPM recordings are associated with all-cause mortality in hypertensive individuals aged 80 and above. This research will help develop more precise and personalized treatment strategies for elderly hypertensive patients, ultimately improving their quality of life and survival rates.

## 2. Methods

### 2.1 Study design and participants

This is a retrospective cohort study that explored whether BPV derived from multiple 24-hour ABPM recordings are associated with all-cause mortality in elderly hypertensive patients. Here, we have 5,838 ABPM records from the Geriatrics Department of Beijing Friendship Hospital, collected from October 12, 2018, to June 9, 2025. These records include the patient’s examination time, patients’ registration number, name, gender, age, and blood pressure measurement results. Patients were searched in the electronic medical record system using their registration number and name, then screened based on their medical record information. The data were filtered according to the following inclusion and exclusion criteria. The inclusion criteria were: (a) aged 80 years or older; (b) complete ABPM recording data; (c) complete clinical data is available. The exclusion criteria included: (a) No history of hypertension; (b) Follow-up period less than six months; (c) Lost to follow-up; (d) Secondary hypertension; (e) Acute cardiovascular events at the time of enrollment. By using the patient’s registration number and name, the number of 24-hour ambulatory blood pressure monitoring sessions completed by each patient during the follow-up period was determined. Based on the number of sessions, patients who completed at least twice ABPM and patients received three or more ambulatory blood pressure measurements during the follow-up period will be included in the subgroup analysis. The entire screening process is shown in Figure 1.

**Figure1.**
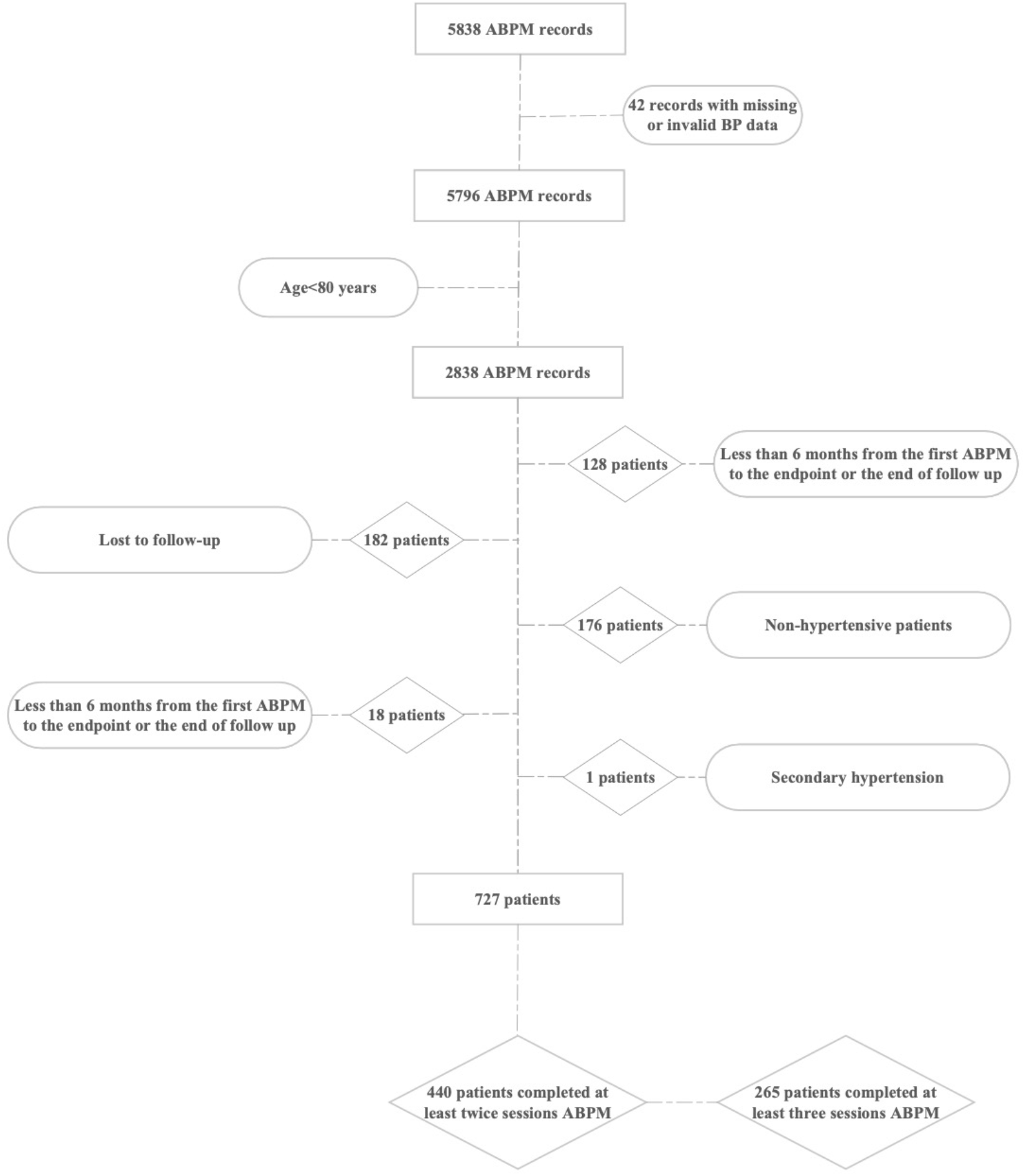
Study flow.

### 2.2 Data collection

Baseline parameters were collected, including demographic information, history of chronic diseases (coronary artery disease [CAD], old cerebral infarction [OCI], diabetes, chronic kidney disease [CKD], atrial fibrillation (AF)), history of smoking, laboratory tests (glycated hemoglobin [HbA1c], high-density lipoprotein cholesterol [HDL-C], low-density lipoprotein cholesterol [LDL-C], creatinine [Cr]) and whether to use antihypertensive drugs. Based on the ABPM results, the following parameters were collected for all patients: 24-hour average systolic blood pressure (24hSBP) and 24-hour average diastolic blood pressure (24hDBP), nighttime systolic blood pressure (nighttime SBP) and diastolic blood pressure (nighttime DBP), and daytime systolic blood pressure (daytime SBP) and diastolic blood pressure (daytime DBP). The nighttime period was defined as 10:00 PM to 6:00 AM, while the remaining hours have been considered daytime. Blood pressure was measured every 30 minutes during the daytime and once every hour during the nighttime. The blood pressure readings at all time points in each patient’s first ABPM report were recorded, along with whether the blood pressure was reverse dipping type. Reverse dipping type was defined as nighttime blood pressure either increasing or decreasing by less than 10% (nighttime blood pressure drop <10%). For patients who have completed two or more ABPM sessions, the 24-hour average systolic and diastolic blood pressure, as well as nighttime and daytime systolic and diastolic values, were recorded for each session.

### 2.3 Assessment of mortality and grouping

Mortality data were extracted from the electronic medical record (EMR) system of Beijing Friendship Hospital, covering patients enrolled between October 12, 2018, to June 9, 2025. Death records were identified through: discharge status marked as “deceased” in EMR. Follow-up ended at the data of death or study cutoff data (June 9, 2025). Patients who meet the inclusion and exclusion criteria will be included in the study and then divided into a death group and a non-death group. Additionally, patients completed twice or more ambulatory blood pressure measurements during the follow-up period will be included in the subgroups analysis. This study was approved by the Ethics Committee of Beijing friendship hospital. The ethic review approval number is 2026-P2-085-01 and the clinical trial number is MR-11-26-021030.

### 2.4 Statistics analysis

All statistical analyses were carried out using Python, continuous variables were reported as mean ±standard deviation (x ±SD) or median (interquartile range) according to whether they were normal distribution. Categorical variables were expressed as frequencies [n, (%)]. Intergroup measurement comparisons were performed using t-tests for normally distributed data or Wilcoxon rank sum tests for non-normally distributed data. Counts were compared by chi-square test.

Based on the first ABPM recordings of all patients who meet the inclusion and exclusion criteria, the blood pressure values at each measurement point have been used to calculate the 24hSBP, 24hDBP, daytime SBP, daytime DBP, night SBP, and night DBP blood pressure variability (BPV). The BPV indicators include the standard deviation (SD), which is the most commonly used and practical variability index, as well as other BPV indicators such as the coefficient of variation (CV) (calculated as SD/mean BP * 100%), average real variability (ARV) (the average absolute variation between consecutive BP measurements) and variation independent of the mean (VIM). VIM was defined as the standard deviation divided by mean x, where x represents the regression coefficient obtained from regressing the natural logarithm of the SD on the natural logarithm of the mean(5). Subsequently, for patients who completed two ABPM sessions, the absolute values of the 24hSBP, 24hDBP, daytime SBP, daytime DBP, nighttime SBP, and nighttime DBP for these two sessions have been calculated. Finally, for patients who completed three or more ABPM sessions, the BPV for 24hSBP, 24hDBP, daytime SBP, daytime DBP, nighttime SBP, and nighttime DBP has been calculated, with the BPV indicators as described above.

Cox regression analysis was employed to examine the relationship between BPV and mortality. Initially, univariate cox analysis was performed to assess their associations, followed by multivariate cox analysis with adjustment for potential risk factors. In our study, variables with p<0.05 in univariate cox regression were included in the multivariate model. Additionally, clinically important covariates such as age, gender, BMI and hypertensive drugs use were forced into the model regardless of their univariate significance. To avoid the impact of multicollinearity on the model results, separate multivariable cox regression analyses were conducted for blood pressure measurements at different time points (e.g., 24hSBP, 24hDBP, daytime SBP, daytime DBP, nighttime SBP, and nighttime DBP) and BPV. In each analysis, potential confounders, including age, sex, BMI, and certain chronic diseases, were controlled for. The results of the multivariate cox analyses were presented using forest plots. Survival rates were estimated using the Kaplan-Meier (KM) method, and differences in survival curves between groups were compared using the log-rank test. P-value < 0.05 was considered statistically significant.

## 3. Results

### 3.1 Participant characteristics

A total of 727 patients were enrolled in the study based on the inclusion and exclusion criteria (Figure 1), Among them, 440 patients underwent at least two ABPM sessions, while 265 patients had complete data from three or more ABPM sessions. In a median follow-up period of 32.0 months (19.0, 50.0), supplementary table 1 described the demographic, clinical, and laboratory test characteristics of all participants at their first ABPM session, stratified by mortality status. A total of 271 patients were assigned to the death group, while 456 patients were assigned to the non-death group. The proportion of reverse dipping type was significantly higher in the death group (52.8%) compared to the non-death group (42.8%), with a P-value of 0.013. As shown in supplementary table 1, the SD value of 24hDBP (9.8±3.6 versus 10.5±3.6, P=0.003) and the SD value of daytime DBP were significantly different between the death and non-death groups. The CV of nighttime DBP was also significantly different between the two groups (11.8±7.2 versus 13.1±7.4, P=0.018). The ARV of 24hSBP, 24hDBP, daytime SBP, and daytime DBP was calculated for both the death and non-death groups. The results showed that the ARV in the death group was 9.5±3.4, 7.0±3.2, 9.4±3.6, and 6.9±3.2, which were significantly lower than the non-death group values of 10.1±3.3, 7.8±3.2, 10.1±3.6, and 7.9±3.5, with P values of 0.016, <0.001, 0.016, and <0.001, respectively. There were also significant differences in VIM between the death group and the non-death group in 24hDBP (9.1 [7.1, 11.3] vs. 9.6 [7.8, 12.7]), daytime DBP (8.8 [6.5, 11.3] vs. 9.4 [7.2, 12.5]), and nighttime DBP (6.8 [4.5, 9.5] vs. 7.4 [5.0, 10.0]), with p-values of 0.001, 0.008, and 0.039, respectively. These results suggest that the 24-hour BPV shows less pronounced characteristics in elderly hypertensive patients in the death group.

Subgroup analyses were conducted for individuals who completed two ABPM sessions and those who completed two or more ABPM sessions. The participant characteristics for these groups are shown in supplementary tables 2 and 3, respectively. Supplementary table 2 shows that a total of 440 participants completed at least two ABPM sessions. The absolute differences between the first and second measurements for 24hSBP, 24hDBP, daytime SBP, daytime DBP, nighttime SBP, and nighttime DBP were calculated and analyzed using non-parametric tests. The results indicated that the difference in nighttime SBP was statistically significant between the death group (11.0 [6.0, 19.5]) and the non-death group (8.0 [4.0, 17.0]), with a p-value of 0.009. No significant differences were observed for the other values, as all p-values were greater than 0.05. Supplementary table 3 demonstrates that 265 participants underwent three or more ABPM sessions. BPV indicators, including SD, CV, ARV and VIM were assessed for 24hSBP, 24hDBP, daytime SBP, daytime DBP, nighttime SBP, and nighttime DBP in both the death and non-death groups. The SD of daytime DBP and nighttime SBP were higher in the death group compared to the non-death group, with values of 6.2±4.3 versus 5.2±2.8 and 12.5±7.1 versus 10.1±5.4, respectively, both showing statistical significance, with P-values of 0.024 and 0.002. The CV analysis showed that 24hDBP, daytime DBP, and nighttime SBP were significantly higher in the death group compared to the non-death group, with p-values of 0.044, 0.008, and 0.007, respectively, indicating statistical significance between the two groups. For the ARV, the median values for daytime DBP and nighttime SBP were 6.3 (4.0, 9.0) and 12.7 (8.5, 17.3) in the death group, significantly higher than those in the non-death group, which were 5.3 (3.7, 7.0) and 10.6 (6.4, 15.1), with p-values of 0.015 and 0.003, respectively. Significant differences in VIM were observed between the death group and the non-death group for 24hDBP (5.3 [3.6,7.5] vs.4.3 [3.1, 15.8]), daytime DBP (5.5 [3.9, 7.3] vs. 4.5 [3.4, 5.9]), and nighttime DBP (9.4 [5.5, 13.9] vs. 5.5[4.1, 7.6]), with P values of 0.015, 0.010, and 0.014, respectively.

### 3.2 Short-term BPV and mortality in elderly hypertensive patients

Univariate cox analysis displayed the reverse-dipper blood pressure pattern was associated with a higher risk of mortality (HR = 1.584, 95% CI: 1.247–2.011, P < 0.001), and this association persisted after adjustment for age, gender, BMI, OCI, AF, CKD, antihypertensive drugs, LDL-C, and HDL-C (HR = 1.489, 95% CI: 1.199–1.850, P < 0.001) (Figure 2a). The KM curves also showed a significantly lower survival rate in the group with a higher prevalence of the reverse-dipper blood pressure pattern (P < 0.001, figure 2b). Regarding short-term BPV, univariate cox analysis demonstrated that multiple 24-hour and daytime BPV indices—including 24hDBP-SD (HR = 0.952, 95% CI: 0.918–0.987, P = 0.007),daytime DBP-SD (HR = 0.950, 95% CI: 0.917–0.984, P = 0.004), 24hSBP-ARV (HR = 0.955, 95% CI: 0.920–0.991, P = 0.013), 24hDBP-ARV (HR = 0.931, 95% CI: 0.893–0.972, P = 0.001), daytime SBP-ARV(HR = 0.956, 95% CI: 0.922–0.991, P = 0.014), daytime DBP-ARV(HR = 0.929, 95% CI: 0.894–0.966, P < 0.001), 24hDBP-VIM (HR = 0.950, 95% CI: 0.9164–0.986, P = 0.007) and daytime DBP-VIM (HR = 0.948, 95% CI: 0.915–0.983, P = 0.004)—were all inversely associated with mortality risk (Table 1). In contrast, short-term nighttime BPV showed no significant association with mortality (all P > 0.05). However, as shown in supplementary figure 1, the significant short-term BPV indicators from the univariate cox analysis did not have a significant impact on mortality after adjustment for age, gender, BMI, OCI, AF, CKD, antihypertensive drugs, LDL-C, and HDL-C.

**Figure 2a.**
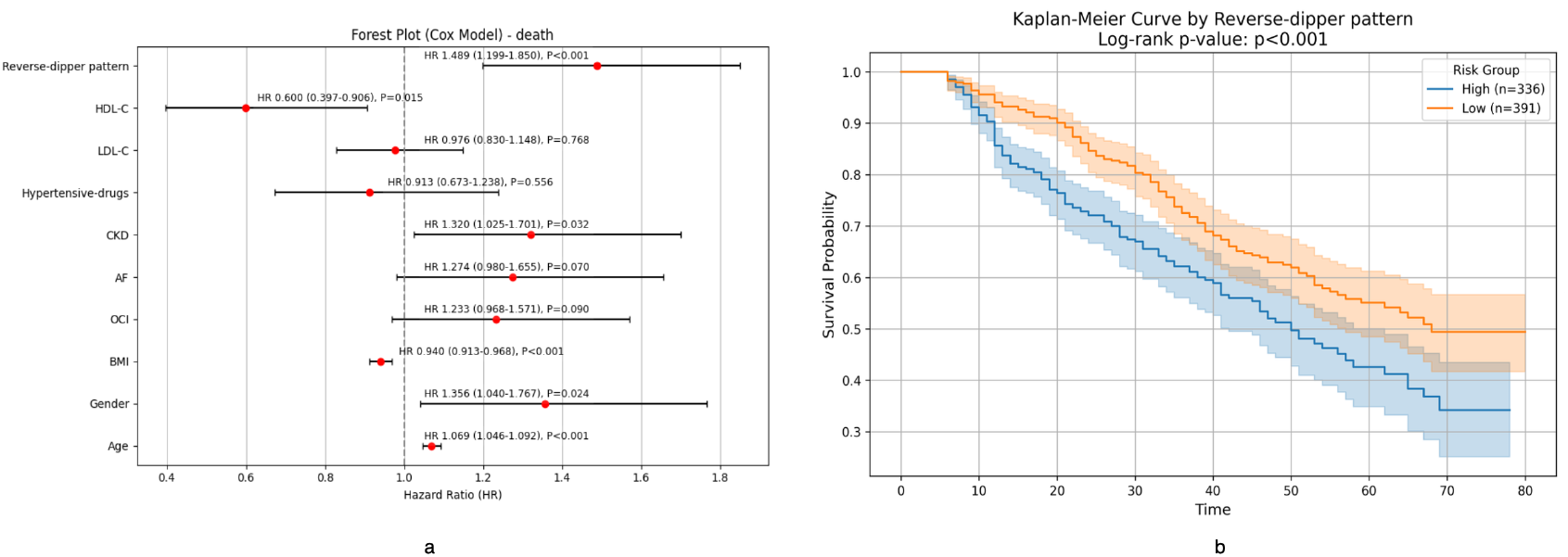
multivariate cox analyses were presented using forest plots showed the reverse-dipper blood pressure pattern was associated with a higher risk of mortality after adjusting for age, gender, BMI, OCI, AF, CKD, hypertensive drugs, LDL-C, and HDL-C (HR = 1.49, 95% CI: 1.20–1.85, P < 0.001). Figure 2b, The KM curves showed a significantly lower survival rate in the group with a higher prevalence of the reverse-dipper blood pressure pattern (log-rank P < 0.001).

**Table 1.** Univariate cox regression analysis for mortality in all patients.

| Parameter | HR | 95%CI | P-value |
| --- | --- | --- | --- |
| Age | 1.098 | 1.073-1.124 | **<0.001** |
| Male | 1.683 | 1.249-2.268 | **<0.001** |
| BMI | 0.938 | 0.910-0.967 | **<0.001** |
| CAD | 1.108 | 0.868-1.413 | 0.409 |
| Diabetes | 0.978 | 0.764-1.251 | 0.857 |
| OCI | 1.537 | 1.186-1.991 | **0.001** |
| AF | 1.446 | 1.094-1.911 | **0.009** |
| CKD | 1.795 | 1.380-2.336 | **<0.001** |
| History of smoking | 1.287 | 0.998-1.660 | 0.052 |
| Hypertensive drugs | 0.830 | 0.601-1.147 | 0.259 |
| HbA1c | 0.953 | 0.859-1.059 | 0.372 |
| LDL-C | 0.819 | 0.685-0.980 | **0.029** |
| HDL-C | 0.401 | 0.253-0.633 | **<0.001** |
| Cr | 1.001 | 1.000-1.003 | 0.108 |
| Daytime DBP | 0.977 | 0.962-0.992 | **0.002** |
| Nighttime SBP | 1.009 | 1.001-1.018 | 0.080 |
| Nighttime DBP | 1.011 | 0.999-1.031 | 0.076 |
| Reverse-dipper pattern | 1.584 | 1.247-2.011 | **<0.001** |
| SD of BP |  |  |  |
| 24h SBP | 0.996 | 0.963-1.029 | 0.802 |
| 24h DBP | 0.952 | 0.918-0.987 | **0.007** |
| Daytime SBP | 0.990 | 0.959-1.023 | 0.548 |
| Daytime DBP | 0.950 | 0.917-0.984 | **0.004** |
| Nighttime SBP | 1.000 | 0.976-1.024 | 0.985 |
| Nighttime DBP | 0.989 | 0.962-1.015 | 0.402 |
| CV of BP |  |  |  |
| 24h SBP | 0.993 | 0.957-1.030 | 0.701 |
| 24h DBP | 0.981 | 0.959-1.003 | 0.092 |
| Daytime SBP | 0.990 | 0.949-1.032 | 0.635 |
| Daytime DBP | 0.979 | 0.957-1.002 | 0.068 |
| Nighttime SBP | 0.989 | 0.958-1.020 | 0.482 |
| Nighttime DBP | 0.984 | 0.967-1.002 | 0.084 |
| ARV of BP |  |  |  |
| 24h SBP | 0.955 | 0.920-0.991 | **0.013** |
| 24h DBP | 0.931 | 0.893-0.972 | **0.001** |
| Daytime SBP | 0.956 | 0.922-0.991 | **0.014** |
| Daytime DBP | 0.929 | 0.894-0.966 | **<0.001** |
| Nighttime SBP | 0.996 | 0.977-1.016 | 0.705 |
| Nighttime DBP | 0.987 | 0.964-1.010 | 0.266 |
| VIM of BP |  |  |  |
| 24h SBP | 0.996 | 0.962-1.030 | 0.801 |
| 24h DBP | 0.950 | 0.916-0.986 | **0.007** |
| Daytime SBP | 0.990 | 0.957-1.023 | 0.542 |
| Daytime DBP | 0.948 | 0.915-0.983 | **0.004** |
| Nighttime SBP | 0.998 | 0.974-1.023 | 0.891 |
| Nighttime DBP | 0.987 | 0.960-1.015 | 0.351 |

### 3.3 Nighttime SBP variability, based on absolute blood pressure differences, was linked to mortality in elderly patients with hypertension

In a median follow-up period of 36.5 months (23,0, 52,0), univariate cox analysis of the absolute differences in 24hSBP, 24hDBP, daytime SBP, daytime DBP, nighttime SBP, and nighttime DBP, calculated from two consecutive ABPM, revealed that higher absolute value of nighttime SBP was closely associated with increased mortality in elderly hypertensive patients (HR=1.019, 95%CI: 1.007-1.031, P=0.002), while the others showed no statistical significance (Supplementary table 3). As shown in supplementary figure 2a, further multivariate cox regression analysis, which was displayed as a forest plot, after adjusting for age, gender, CKD, OCI, hypertension drugs, and HDL-C, still showed a significant association between nighttime SBP variability and mortality (HR=1.014, 95%CI:1.003-1026, P=0.010). KM survival analysis was performed to evaluate the association between nighttime SBP variability and mortality in elderly hypertensive patients. Participants were divided into two groups based on median nighttime SBP variability: high variability group and low variability group. The KM curves demonstrated that the high variability group had significantly lower survival rates over the 36.5 months (23,0, 52,0) follow-up period. The log-rank test revealed a statistically significant difference between the groups (P=0.010, supplementary figure 2b).

### 3.4 Long-term BPV and mortality in elderly hypertensive patients

Table 2 presents the results of univariate cox analysis for BPV, calculated from three or more ABPM measurements, and its relationship with mortality, with a follow-up duration of 41.0 (28.0, 56.0) months. The analysis reveals that higher SD, CV, ARV and VIM of daytime DBP (HR=1.048, 95%CI:1.007-1.090, P=0.024, HR=1.043, 95%CI:1.011-1.077, P=0.009, HR=1.032, 95%CI:1.010-1.055, P=0.005, HR=1.049, 95%CI:1.005-1.094, P=0.027, respectively), as well as higher SD, CV, ARV and VIM of nighttime SBP (HR=1.046, 95%CI:1.018-1.075, P=0.001, HR=1.056, 95%CI:1.018-1.095, P=0.003, HR=1.039, 95%CI:1.021-1.057, p<0.001, HR=1.048, 95%CI:1.019-1.078, p=0.001, respectively), all demonstrated significant differences. Moreover, an increased ARV of nighttime DBP and (HR=1.027, 95%CI:1.003-1.052, P=0.025) was also significantly associated with mortality. Further multivariate cox regression analysis revealed that after adjusting for age, gender, BMI, smoking history, CKD, antihypertensive drugs use, OCI, and AF, several BPV-related indicators, such as CV of daytime DBP (HR=1.032, 95%CI: 1.003-1.062, P=0.033), and SD, CV, ARV and of nighttime SBP (HR=1.036, 95%CI:1.009-1.063, P=0.007, HR=1.039, 95%CI:1.005-1.075, P=0.026, HR=1.031, 95%CI:1.014-1.048, p<0.001, HR=1.037, 95%CI:1.010-1.065, p=0.008, respectively), all showed significant associations with mortality, which was displayed as a forest plot (Figure 3). This indicates that higher BPV is significantly associated with mortality in elderly hypertensive patients. KM survival analysis were used to evaluate the relationship between long-term blood pressure variability and mortality in elderly hypertensive patients. The results showed that during a follow-up period of 41.0 months (28.0, 56.0), patients with high daytime DBP-CV and high nighttime SBP-ARV had significantly lower survival rates. Log-rank test revealed statistically significant differences (P=0.012, P=0.024, respectively, Figure 4).

**Table 2.** Univariate cox analysis of BPV for mortality in patients who completed a minimum of three sessions of ABPM.

| Parameter | HR | 95%CI | P-value |
| --- | --- | --- | --- |
| Age | 1.104 | 1.068-1.142 | **<0.001** |
| Male | 1.623 | 1.056-2.493 | **0.027** |
| BMI | 0.892 | 0.846-0.941 | **<0.001** |
| CAD | 1.168 | 0.084-1.695 | 0.415 |
| Diabetes | 1.036 | 0.717-1.496 | 0.850 |
| OCI | 1.543 | 1.037-2.298 | **0.033** |
| AF | 1.518 | 1.015-2.271 | **0.042** |
| CKD | 2.016 | 1.383-2.937 | **<0.001** |
| History of smoking (n, %) | 1.752 | 1.210-2.536 | **0.003** |
| Hypertensive drugs | 0.688 | 0.421-1.124 | 0.135 |
| HbA1c (%) | 0.870 | 0.720-1.051 | 0.148 |
| LDL-C | 0.890 | 0.694-1.141 | 0.357 |
| HDL-C | 0.590 | 0.313-1.111 | 0.102 |
| Cr | 1.001 | 0.999-1.003 | 0.304 |
| SD of BP |  |  |  |
| 24h SBP | 1.024 | 0.984-1.065 | 0.246 |
| 24h DBP | 1.021 | 0.973-1.072 | 0.403 |
| Daytime SBP | 1.011 | 0.979-1.044 | 0.505 |
| Daytime DBP | 1.048 | 1.007-1.090 | **0.024** |
| Nighttime SBP | 1.046 | 1.018-1.075 | **0.001** |
| Nighttime DBP | 1.035 | 0.994-1.078 | 0.096 |
| CV of BP (%) |  |  |  |
| 24h SBP | 1.030 | 0.978-1.085 | 0.263 |
| 24h DBP | 1.028 | 0.990-1.067 | 0.152 |
| Daytime SBP | 1.012 | 0.973-1.054 | 0.545 |
| Daytime DBP | 1.043 | 1.011-1.077 | **0.009** |
| Nighttime SBP | 1.056 | 1.018-1.095 | **0.003** |
| Nighttime DBP | 1.022 | 0.990-1.056 | 0.174 |
| ARV of BP |  |  |  |
| 24h SBP | 1.030 | 0.996-1.064 | 0.082 |
| 24h DBP | 1.035 | 0.992-1.080 | 0.113 |
| Daytime SBP | 1.020 | 0.994-1.047 | 0.136 |
| Daytime DBP | 1.032 | 1.010-1.055 | **0.005** |
| Nighttime SBP | 1.039 | 1.021-1.057 | **<0.001** |
| Nighttime DBP | 1.027 | 1.003-1.052 | **0.025** |
| VIM of BP |  |  |  |
| 24h SBP | 1.024 | 0.984-1.067 | 0.242 |
| 24h DBP | 1.022 | 0.972-1.074 | 0.398 |
| Daytime SBP | 1.011 | 0.979-1.045 | 0.502 |
| Daytime DBP | 1.049 | 1.005-1.094 | **0.027** |
| Nighttime SBP | 1.048 | 1.019-1.078 | **0.001** |
| Nighttime DBP | 1.037 | 0.994-1.081 | 0.093 |

**Figure 3.**
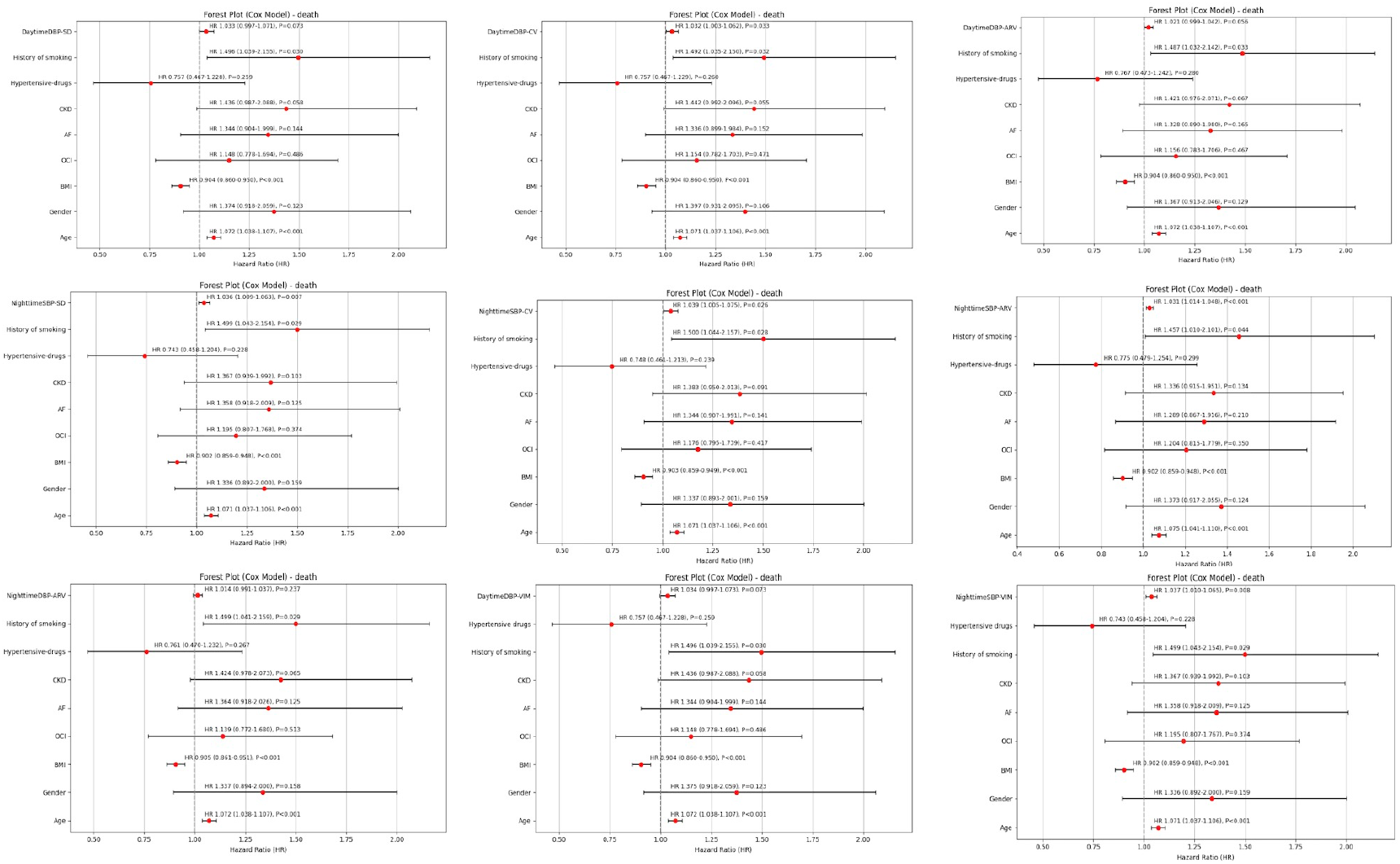
Multivariate Cox analysis, after adjusting for age, gender, BMI, OCI, AF, CKD, antihypertensive drugs, and smoking history, revealed that daytime DBP-SD, daytime DBP-ARV, and nighttime DBP-ARV did not significantly affect mortality. In contrast, daytime DBP-CV, nighttime SBP-SD, nighttime SBP-CV, nighttime SBP-ARV and nighttime SBP-VIM were found to have a significant impact on mortality.

**Figure 4.**
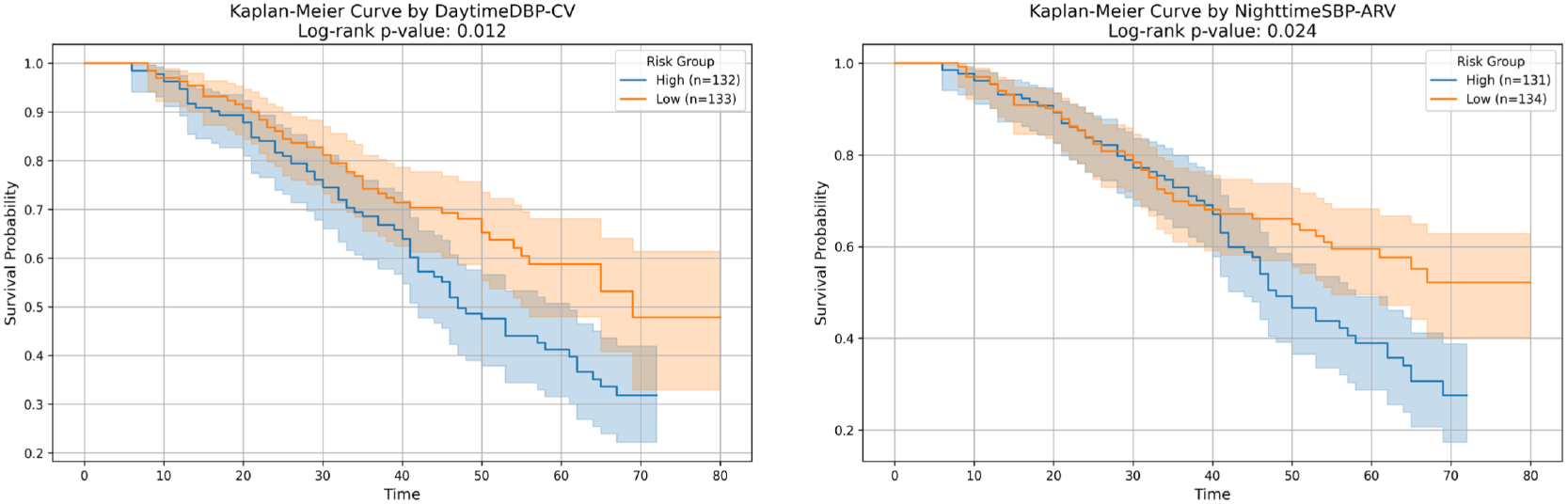
KM curves demonstrated significantly lower survival rates in patients with higher daytime DBP-CV and increased nighttime SBP-ARV (log-rank P=0.012, p=0.024, respectively).

## 4. Discussion

Our study focused on elderly hypertensive patients aged 80 and above and conducted a retrospective analysis to assess the impact of blood pressure measurements through ABPM on mortality. Circadian variation in blood pressure plays a crucial role in cardiovascular health and patient outcomes. Reverse dippers — those with higher nocturnal blood pressure compared to daytime blood pressure — are known to have an increased cardiovascular risk, more frequent hypertension-mediated organ damage, and higher mortality than normal dippers (6-8). Reverse dipping is especially prevalent among elderly hypertensive patients. While many studies have focused on patients with an average age over 60, few have specifically examined those aged 80 and above. Our study, however, specifically targets this age group. We observed that reverse dipping blood pressure was significantly more common in the death group compared to the non-death group (52.2% vs. 42.8%, P=0.013). Univariate cox analysis and multivariate cox analysis adjusted for various covariates confirmed the negative impact of reverse dipping blood pressure on mortality in elderly hypertensive patients. The difference in KM survival curves between the two groups with reverse dipping blood pressure was significant, with the mortality group showing a significantly higher percentage of reverse dipping blood pressure. It suggests that reverse dipping blood pressure may serve as an independent clinical factor indicating a higher risk of mortality. This further emphasizes that in elderly hypertensive patients, abnormalities in blood pressure circadian rhythms could be a key factor in assessing long-term prognosis.

In this study, we calculated the impact of short-term BPV from the first dynamic blood pressure measurement, as well as long-term BPV, on mortality. Univariate cox analysis found a negative correlation between higher BPV within 24 hours and mortality risk, which is inconsistent with some previous studies(9, 10). However, this significant association disappeared after adjusting for multiple risk factors, suggesting that the relationship between short-term BPV and mortality risk may be influenced by these covariates. Our study differs from previous research in that it included only elderly patients aged 80 and above, and this discrepancy may also indicate that smaller fluctuations in 24-hour BPV may not always directly reflect a healthy physiological state. In elderly hypertensive patients, this may reflect compromised hemodynamic regulation or impaired sympathetic nervous function. However, in the multivariate model, this effect could be confounded or accounted for by other established mortality risk factors, which may explain the lack of an independent association. Consequently, further studies are warranted to better elucidate this relationship.

Studies have shown that BPV, rather than average BP, is linked to adverse cardiovascular events, with the majority of cardiovascular events in the modern era occurring in individuals with blood pressure under 140/90 mmHg(11-13). This analysis suggests that BPV could be a key factor in determining outcomes for high risk hypertensive patients, even when blood pressure is well controlled, underscoring the need to focus on both blood pressure stability and numerical targets for optimal risk reduction(14). Over time, the calculation of BPV from multiple ABPM measurements in our study showed that greater long term BPV, whether during the day or night, was significantly associated with mortality risk. This is consistent with several previous studies. Our research particularly emphasizes the importance of nighttime BPV. Both the absolute changes in nighttime SBP between consecutive measurements and the BPV of nighttime SBP derived from three or more ABPM indicate a close association between high nighttime BPV and mortality. A previous study(15) with an average age of 58 found that higher BPV, particularly systolic BPV, was closely associated with clinical outcomes. This study suggested that the stronger association with systolic BPV may be linked to the relatively older age of participants included. Our study, which included patients with a higher average age, further underscores the increasing prognostic significance of systolic blood pressure with age.

Compared with office or home blood pressure measurements, ABPM provides a more comprehensive and physiologically relevant assessment of long-term blood pressure variability by capturing 24-hour, daytime, and nighttime fluctuations, while minimizing measurement bias. In a comprehensive study of both clinic and ambulatory blood pressure, ambulatory blood pressure monitoring offered more significant insights into the risk of all-cause mortality and cardiovascular death than traditional clinic blood pressure measurements(16). This advantage is particularly important in elderly populations, in whom abnormal nocturnal blood pressure patterns and increased BPV are more prevalent and clinically meaningful. As our study found, long-term variability in nighttime SBP is very common in elderly hypertensive patients. Multivariate cox analysis revealed that higher nighttime SBP variability was significantly associated with higher mortality. This variability was assessed using SD, CV ARV and VIM, all of which were significantly higher in the mortality group. Furthermore, the KM survival curves further supported the cox regression findings, demonstrating a markedly lower survival probability among patients with higher nighttime SBP variability, suggesting its potential prognostic significance. Prior research has shown that nighttime BPV is linked to target organ damage and may be associated with an increased risk of mortality(16, 17). Research has also indicated that elevated nighttime BP is linked to target organ damage, such as chronic kidney disease and heart failure, which may contribute to an increased risk of mortality(18, 19). Other previous studies have also observed the superiority of nighttime blood pressure over daytime blood pressure(20-23). However, in the present study, long-term higher daytime DBP-CV was independently associated with increased mortality in elderly hypertensive patients. Daytime DBP variability may reflect impaired vascular compliance and unstable peripheral vascular resistance, which are common features in advanced aging. ABPM-based assessment revealed that long-term variability in nighttime SBP and daytime DBP is independently associated with increased mortality in elderly hypertensive patients, underscoring the prognostic value of BPV and circadian patterns beyond absolute blood pressure levels.

This study has some limitations. first, the data is derived from a single center, and the study results may not be widely applicable to other regions or different populations. Future studies should adopt multicenter designs to further validate these findings and enhance the generalizability of the results. Second, although medication data were collected at baseline, information on medication use was not available during the follow-up period. Third, the current study does not examine the association between ABPM and non-fatal events, as only death data were available.

In conclusion, in this retrospective analysis of ABPM data from individuals aged 80 years and older, the reverse dipper blood pressure pattern was independently associated with an increased risk of death. While short-term blood pressure variability was not related to mortality, greater long-term variability in nighttime SBP and daytime DBP were strongly associated with increased mortality. These findings highlight the importance of blood pressure patterns and long-term stability, beyond absolute blood pressure levels, in very elderly hypertensive patients. Our findings emphasize that achieving blood pressure stability and preserving normal circadian patterns may be important considerations in the management of hypertension in very elderly individuals.

## Acknowledgements

None.

## Authors’ contributes

Mengying Zeng and Ying Sun designed the experiments. Mengying Zeng, Mengjun Jiang, Yuanyuan Shang, Jingwen Shi and and Yabing, Wang were responsible for data collection and organization. Mengying Zeng and Yuchen Zhu performed the experiments and data analysis. Mengying Zeng and Ying Sun wrote the manuscript. All authors read and approved the final manuscript.

## Data availability

The datasets generated and analyzed during the current study are not publicly available due the specificity of the personnel and the confidentiality of the data. But are available from the corresponding author on reasonable request.

## Declarations

### Ethics approval and consent to participate

This study was approved by the Institutional Review Board (IRB) of Beijing Friendship Hospital, and informed consent was obtained from all participants. The research adhered to the principles of the Declaration of Helsinki. All data were anonymized, and participant confidentiality was ensured throughout the study.”

### Consent for publication

Not applicable.

